# Motor Improvement in Neurological Conditions (MINC): Multiple Sclerosis Design and methods of a single-arm feasibility study

**DOI:** 10.1101/2023.07.30.23293287

**Authors:** Onno van der Groen, Yvonne Learmonth, Kirsten van Rijn, James Smith, Dylan Edwards

## Abstract

**Introduction:** Multiple sclerosis (MS) is a chronic progressive neurological disease. There is ample evidence that exercise can be beneficial. The advancement of modern technology led to improvements in the way therapy can be offered and can make it more motivating, thereby increasing adherence. The primary objective of this two site single blinded randomized control trial (RCT) is to explore the feasibility of conducting a multicentre definite RCT trial with a neuroanimation intervention of high-dose practice in people with mild-to-moderate MS. The secondary objective is to collect data on the variability of outcome measures to inform sample size calculations for a RCT. The tertiary outcome is to assess if this intervention changes exercise behaviour.

**Methods and analysis:** This study is in preparation for a future definitive randomised control trial (RCT) where the efficacy compared to a dose matched control therapy will be assessed. The setting for this study is a research laboratory at Edith Cowan University (ECU) and a neurological service provider, Multiple Sclerosis Society of Western Australia (MSWA). This feasibility study will recruit people with MS who have mild to moderate disability. Subjects will participate in 24 session, 2 times a week, of 60 minutes time-on-task intense arm training, using an exergaming system. Participants will undergo a follow up within 3 days and at 6 months after the final study visit.

**Ethics and dissemination:** This study was approved by the local Ethics Committee of Edith Cowan University. Subjects will be included after signing informed consent. Study outcomes will be disseminated through presentations at scientific conferences and through peer-reviewed journals.

**Trial registration:** ACTRN12622000281796

**Strengths and limitations of this study:**

- The study intervention is a newly developed exercise intervention protocol designed to be engaging and motivating
- Next to investigating primary efficacy in order to determine sample size for a larger trial, the study also uses implementation science to assess future obstacles in a follow up randomized control trial
- The feasibility of conducting a larger trial will be based on standardised criteria regarding process, resource, and management metrics
- This study without a control group demonstrates feasibility rather than efficacy

## 1. Introduction

Physical activity provides many benefits for people with multiple sclerosis (MS), including improvements in physical fitness, mobility, muscle strength as well as possible secondary benefits such as improved quality of life, mood and well-being [1–3]. People with MS are typically less active than the general population [4,5]. There are established guidelines for exercise in persons with MS [6,7], in addition to resistance training, aerobic exercise recommendations are 2 – 3 days/week of aerobic training (10 – 30 minutes of moderate-intensity). Adhering to these guidelines might reduce fatigue, improve mobility, and improve quality of life [8]. Several barriers are in place which can prevent people adhering to these guidelines, including physical (e.g., fatigue, pain), psychological (e.g., depression) and environmental factors (e.g., lack of exercise opportunity) [9]. Making exercise a more enjoyable experience could optimise exercise behaviour and lead to better adherence [9,10]. The advancement of modern technology led to improvements in the way therapy can get offered. Exergames involves the integration of physical activity into a video game environment that requires active body movements to control the in-game experience. There is evidence that exergames can be as good as usual physical therapy with comparable dose efficacy but are often more motivating and engaging than conventional rehabilitation programs for individuals with neurological disability [11,12]. Qualitative research demonstrated that exergaming in MS can be beneficial [13] and is experienced as enjoyable by people with MS and treating physiotherapists [14,15]. Moreover, exergame based rehabilitation intervention performed in a health care setting led to statistically significant improvements in health-related quality of life in persons with chronic disease [16].

In this study we will use an immersive exergaming intervention, the MindPod (MindMaze Inc.) motion capture and gaming system. 3D movements of the arm control the movement of a virtual dolphin, swimming through different ocean scenes with various task goals including chasing and eating fish, eluding attacks, and performing jumps. Tasks are designed to promote movement of the upper body in all planes throughout the active ranges of motion and titrated based on successful completion of progressive levels of difficulty (Krakauer et al., 2021). The MindPod can support high dose practice, which is cognitively engaging and has been associated with promising improvements in sensory motor upper-limb function post stroke (Krakauer et al., 2021). However, the feasibility of this intense exergaming protocol has not been established for MS.

## Aims and objectives

### Primary objective

Explore the feasibility of conducting a multicentre definite randomized control trial (RCT) with a neuro-animation intervention system.

### Secondary objective

Collect data on the variability of outcome measures to inform sample size calculations for a larger RCT.

### Tertiary objective

As a tertiary objective we assess if this intervention changes exercise behaviours.

## 2. Methods and analysis

### Study design

The Motor Improvement In Neurological Conditions (MINC): Multiple Sclerosis trial is designed to test the feasibility of a high-dose, high-intensity exergaming trial. This is a two-sites, single arm prepost intervention. The intervention is 12 weeks and participants train twice per week aiming for an hour time-on-task. Participants will train for 24 sessions, either at ECU Joondalup or MSWA Wilson. Participants will be asked to complete 3 assessments visits, that is at baseline, within 3 days and at 6 months of the last training session.

Participants will be asked to visit ECU Joondalup for the assessments where possible. See table 2 for an overview of the assessments collected at each time point.

### Participant characteristics and eligibility

Participants who meet the following criteria will be included: (a) at least 18 years old ; (b) Clinically definite diagnosis of MS ; (c) Patient determined disease step (PDDS) >0 and <= 3(measure of MS severity); (d) Stable medical treatment at least 1 month prior to intervention; (e) not already participating in exercise research; (f) no diagnosis of another neurological illness or musculoskeletal disorder different to MS; (g) no diagnosis of a cardiovascular, respiratory, or metabolic illness or other conditions which may interfere with the study; (h) no flare-up, relapse or hospitalization in the last month prior to commencement of the assessment protocol or during the process of the therapeutic intervention; (i) not pregnant; (j) visual impairment is corrected. These criteria were selected to target people with MS who likely will benefit from this intervention or to protect those for experiencing adverse events.

### Participant recruitment and screening

Between April 2022 and December 2023 people with MS will be recruited through a preexisting observational study (the Systematic Profiling in Neurological Conditions (SPIN) Observational Study), as well as via non-governmental organisations (NGOs), local hospitals and rehabilitation centres, flyers and community advertisements. The study will also be promoted online, that is on websites of relevant health advocacy organisations, and via traditional media, so that interested people with MS can self-refer by contacting the coordinating investigator. The study information letter will be shared with interested potential participants. Interested participants register their interest in this study with the researcher by phone or email. A study researcher will phone those who have expressed an interest in the research to describe the research in more detail, answer questions and go through the inand exclusion criteria. Individuals who fulfil the study criteria will be invited for a screening visit where written consent will be obtained.

### Sample size consideration

The primary aim is to assess feasibility of a high-dose exergaming intervention. In this exploratory study we will determine safety and feasibility in a small group, the sample number will be maximised, however will be limited by available time for study duration and financial resources. We sought a sample size exceeding 12 subjects, as this has deemed acceptable for pilot and feasibility studies [17].

### Intervention

Participants will use the MindPod for up to 60 minutes per session time on task, two times a week for 12 weeks. The exercise frequency of twice per week is based on consumer input. Participants can take breaks if required, which will be logged (measured with a stopwatch) and reported in a logbook. Participants will play 30mins time-on-task with their left arm, and 30mins of time-on-task with the right arm. 3D movements of the arm control the movement of a virtual dolphin, swimming through different oceanic scenes with various goals including chasing and eating fish, eluding attacks, and performing jumps (figure 1).

**Figure 1.**
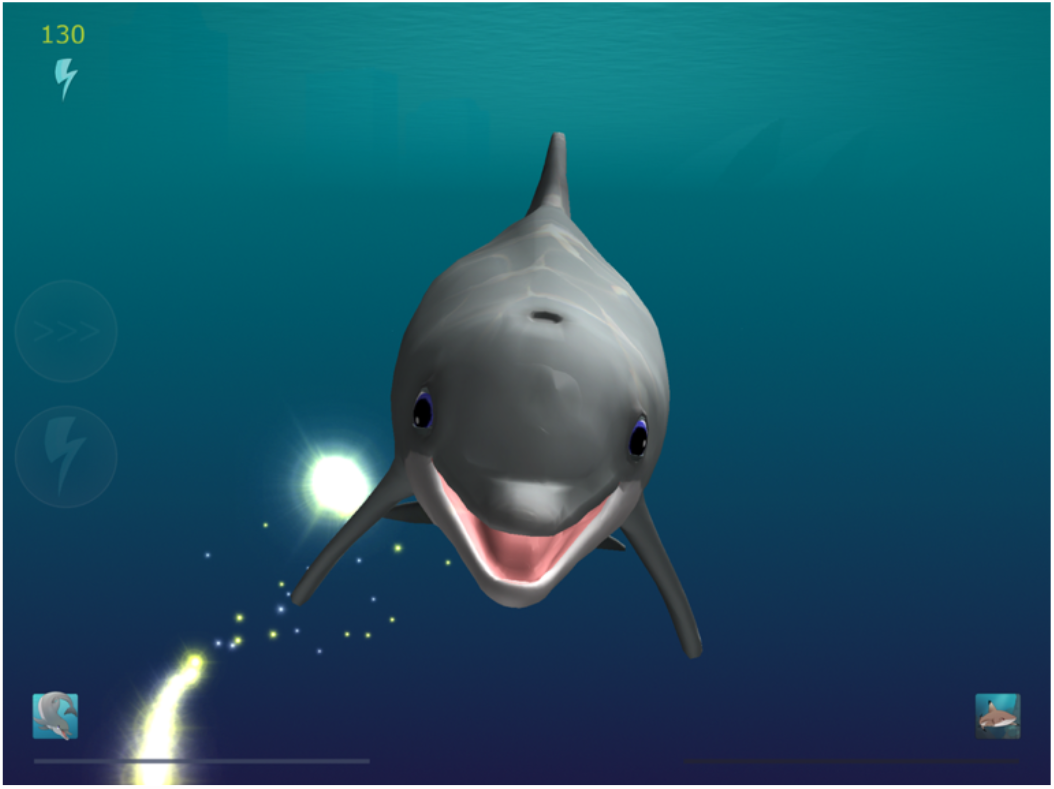
Participants play the MindPod Dolphin game. 3D movements of the arm control the movement of a virtual dolphin, swimming through different ocean scenes with various task goals including chasing and eating fish, eluding attacks, and performing jumps. Tasks are designed to promote movement of the upper body in all planes throughout the active ranges of motion and titrated based on successful completion of progressive levels of difficulty.

Tasks are designed to make movement in all planes throughout active range of motion and titrated based on successful completion of progressive levels of difficulty. A member of the research team will be present throughout each session to supervise. During each session we will determine fatigue levels with a visual analogue scale (VAS). If participants feel fatigued, classified on a 10-point as a visual analogue scale of 8 or higher, they will be offered an antigravity vest (Ekso Bionics, Richmond, CA) to allow participants to exercise for 60 minutes despite excessive fatigue (figure 2). The supervisor adjusts the level of support such that the target rate of perceived exertion (RPE) at the end of each session ranges between 5 and 8. The level of support can be adjusted by either changing the stiffness of the springs in the exoskeleton or removing the exoskeleton altogether.

**Figure 2:**
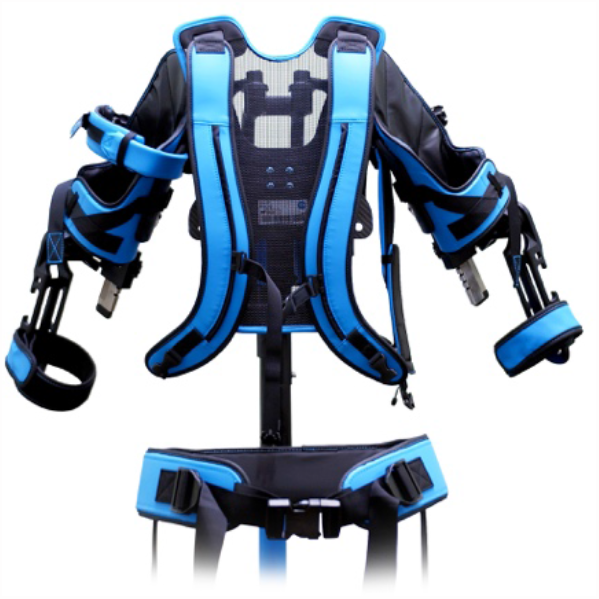
The exoskeleton (Ekso UE, Esko Bionics) is a spring-operated anti-gravity vest. The springs come in different stiffnesses which provide a range of level of support.

### Outcome measures

Since this is a feasibility study, a wide range of outcomes has been included. A detailed description of outcome measures is included in the supplements.

Outcomes will be assessed at baseline, within 3 days and at 6 months after the last training session (figure 3). Clinical assessments are conducted by a qualified physiotherapist or trained clinical research staff, not involved in delivery of the intervention. Self-reported questionnaires will be completed at the person’s own convenience and pace.

**Figure 3:**
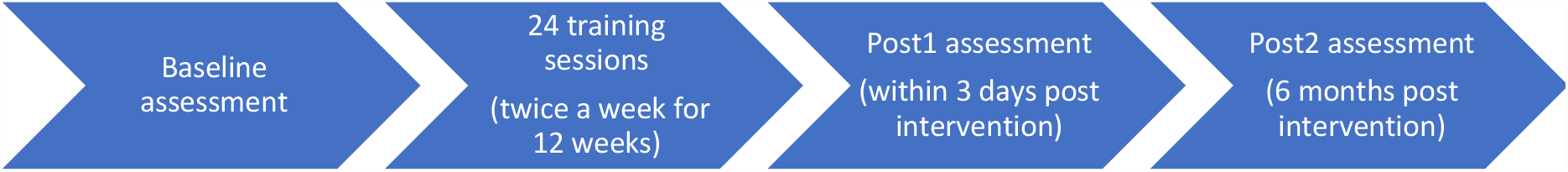
Study assessment timeline. Participants will have one baseline assessment, followed by 24 training sessions of 60min time on task. Post assessments occur within 3 days and at 6 months after the last training session.

### Demographics

The demographics measures include age, gender, occupation, self-identified ethnicity, MS diagnosis date, MS type at diagnosis, any disease modifying therapies, and any symptomatic therapies taken. Note that many trials of exercise training do not collect information around disease modifying therapies[3], therefore there is no clear understanding of the benefits of exercise in the context of disease-modifying drug use.

### Genetic markers

This study is part of a larger observational research program (systematic profiling in neurological conditions, SPIN) where saliva samples will be collected for future analysis. Therefore, participants have the option to provide a saliva sample by passively drooling into a special collection tube at baseline and following the trial to collect their genetic material (DNA and RNA) for future exploratory analyses. New markers or methods for measuring the response to interventions may become available in the future. This would provide invaluable information into the genetic factors that determine whether someone will or will not respond to an intervention.

### Adverse events

Adverse events (AE) are defined as any undesirable outcome, such as injury, falls, discomfort, and pain. All AEs and their possible relation to the intervention will be closely monitored, documented, and reported to the SPIN data Safety Monitoring Committee. The committee will consider the events and offer advice to the project team. The chief investigator will assess all AEs and determine if it should be classified as serious adverse event (SAE). If it is not a SAE then it will be recorded in the study file and the participant will be followed up by the research team. Reports of SAEs will be submitted to the Research Ethics Committee.

### Primary outcome: Feasibility assessments

The primary objective of this study is to assess the feasibility of implementing our intervention for individuals with MS. Therefore, this study will gather outcomes based on process, resource, management and scientific metrics for feasibility, as reported previously [18,19]. These include the evaluation of: i) process: assessing the feasibility of the processes that are key to the success of the main study; ii) resources: assesses the time and resource problems that can occur during the main study; iii) management: assesses potential human and data management problems; iv) scientific: assesses the safety, burden of data collection and response to the study; v) acceptability: the acceptability of this intervention by participants and staff delivering the intervention. In this study, we use Smith’s approach on qualitative proforma development and application in exergames [20]. The Rapid Health Implementation Proforma’s (RHIP) is a rapid qualitative data collection tool used in exergames taking the form of an open-ended, text-box survey, comprising a brief number of questions. RHIP allows participants to expand on survey responses and diversify, to an extent, from the topic area. The metrics are summarised in table 1. The questions asked to evaluate the primary aim are summarised in supplement 2. Participants have the option to answer the questions during a face-to-face interview at the first post-intervention assessment, or during a tele-conference call. The interview will be recorded and transcribed. These questions will be reviewed by a person living with MS

**Table 1:** feasibility metrics of interest.

| Domain | Feasibility objective |
| --- | --- |
| Process | Determine recruitment rates, eligibility, refusal, retention and attrition and adherence, including percentage interested in study after information and screening and reason for attrition |
|  | Determine appropriate eligibility criteria |
|  | Estimate barriers/refusals to participation |
| Resources | Determine the administrative and financial resources needed for the study |
|  | Estimate costs of equipment, space, personal time |
|  | Determine the duration and frequency of communication between participants and staff (email, face-to-face and telephone contact) |
|  | Determine clinical training needs and competence |
| Management | Staffing requirements |
|  | Estimate challenges perceived by study personnel |
|  | Processing time for data collection, data entry and data checking |
|  | Estimate data completeness |
| Scientific | Determine safety |
|  | Determine adverse events |
|  | Compliance |
|  | Treatment effect, eg. data collected on the variability of outcome measures to inform sample size calculations |
|  | Determine participant and therapist reaction to data collection and outcome assessments |
|  | Data collected on if this intervention changes exercise behaviours |
| Acceptability | How an individual feels about the intervention (Affective attitude) |
|  | Determine the perceived amount of effort that is required to participate in the intervention (Burden) |
|  | Determine the extent to which the intervention has a good fit with individual's value system (Ethically) |
|  | Determine the extent to which benefits, profits or values must be given up engaging in the intervention (Opportunity costs) |
|  | Determine the extent to which the participant understands the intervention and how it works (Intervention coherence) |
|  | Determine the extent to which the intervention is perceived as likely to achieve its purpose (Perceived effectiveness) |
|  | Determine the participant's confidence that they can perform the behaviour(s) required to participate in the intervention (Self-efficacy) |

The following minimum success criteria will be used to assess the feasibility of a larger trial[19]:

- No reports of serious adverse events as a result of the intervention
- A minimum of 80% of participants reporting satisfaction
- A minimum of 20% attrition level
- A minimum of 75% recruitment of the intended sample size
- A minimum of 75% completion of each outcome measure

### Secondary outcome

Effect on motor and cognitive function for sample size calculation

This study will gather data on the variability of outcome measures to inform sample size calculations for a future RCT (table 2). Outcome measures will assess different domains, including cognitive functioning, motor functioning, fatigue levels and self-reported quality-oflife measures. Besides this, we collect heart-rate data during each training sessions to determine exercise intensity. After each session we will determine the level of fatigue with a visual analogue scale (VAS). KinArm motor measures are optional for the participants who are able to travel to access the robot necessary to run these tests.

**Table 2:** outcomes measures for sample size calculations.

|  | After each session | Baseline | Within 3 days of the last session | 6 months follow up |
| --- | --- | --- | --- | --- |
| Qualitative questions |  |  | X |  |
| HR-data | X |  |  |  |
| Depression Anxiety Stress Scale |  | X | X | X |
| Symbol Digits Modality Test |  | X | X | X |
| KinArm cognitive tests<br>-Spatial span<br>-Trail making |  | X | X | X |
| Action research arm test (ARAT) |  | X | X | X |
| Fatigue severity scale |  | X | X | X |
| Fatigue visual analogue scale | X |  |  |  |
| Rate of perceived exertion (RPE) | X |  |  |  |
| Timed-up-and go |  | X | X | X |
| Functional walk test |  | X | X | X |
| KinArm motor assessments |  | X | X | X |
| Activities Balance Confidence Scale |  | X | X |  |
| MSWS-12 (MS walking scale 12 self-reported items on walking confidence) |  | X | X | X |
| Leeds MS Quality of life scale |  | X | X | X |
| Saliva (DNA) |  | X |  |  |
| Saliva (RNA) |  | X | X |  |
| Intrinsic Motivation Inventory (IMI) |  | X | X |  |
| Physical Activity Enjoyment Scale (PACES) |  | X | X |  |

### Tertiary outcome: Effect on exercise behaviour

The tertiary outcome assesses the effects of this intervention on exercise behaviour (table 3).

**Table 3:**
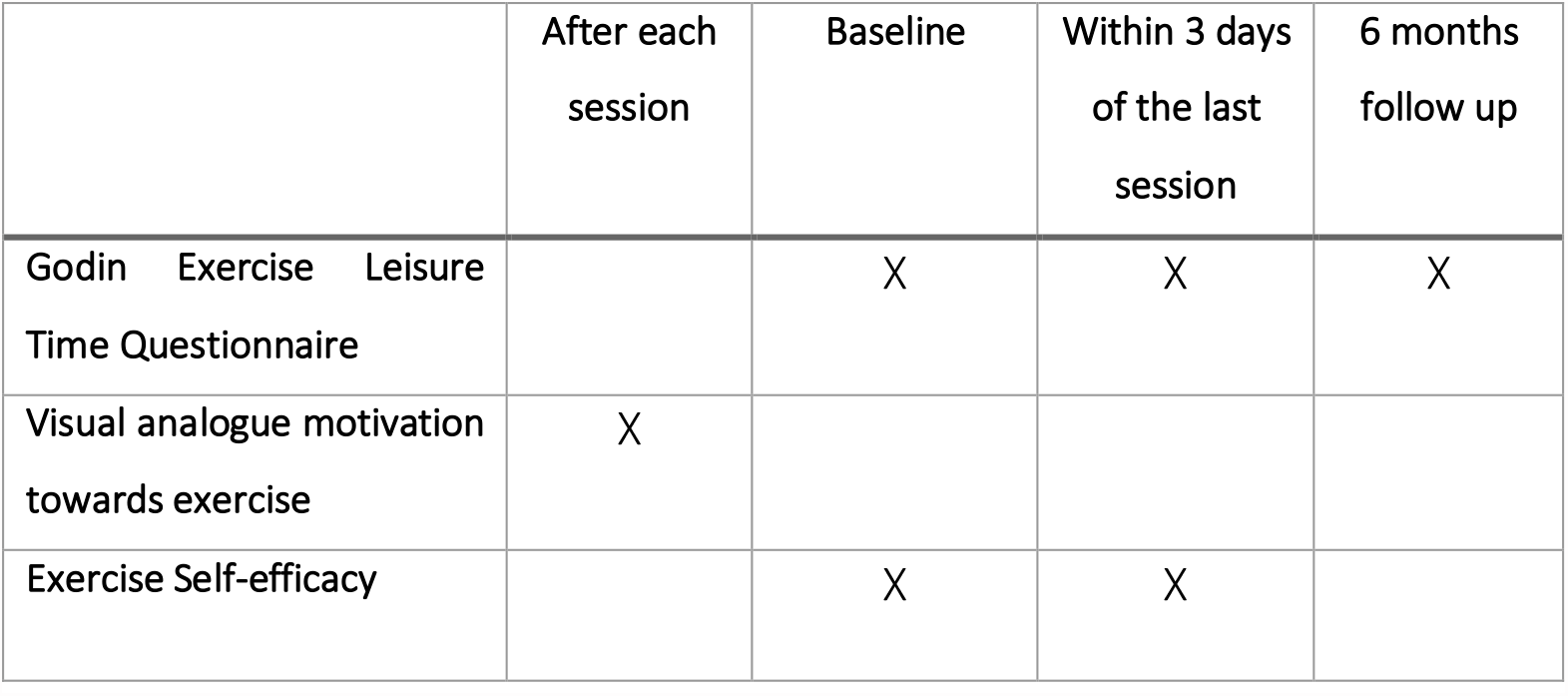
outcomes measures to determine the effects on exercise behaviour.

|  | After each session | Baseline | Within 3 days of the last session | 6 months follow up |
| --- | --- | --- | --- | --- |
| Godin Exercise Leisure Time Questionnaire |  | X | X | X |
| Visual analogue motivation towards exercise | X |  |  |  |
| Exercise Self-efficacy |  | X | X |  |

### Data management

Each participant will receive an unique identifying code which will be recorded on all datasets pertaining to that individual. The coding system will be held in a file separate from the other datasets. All datasets will be stored on a secure research storage platform. On completion data will be stored for 25 years in the ECU institutional repository.

### Data analysis

Data analysis will occur in accordance with the objectives of this study: assessing the feasibility of the current intervention, determine the effects on motor and cognitive function for sample size calculation and its effects on exercise behaviour. Descriptive statistics will be used to report on the feasibility measures, which include process, resources, management and scientific. The process data will inform the feasibility of achieving the required sample size for a larger trial. The resource data will be used to determine the costs of a larger trial. The managment data will be used to determine the administrative requirements for a larget tiral and the scientific domain will identify the suitability of the outcome measures and any risk management for a future trial. A qualitative approach will be used for free text response questions (table 3), with qualitative data analysed using Framework Analysis [21]. Data management will be facilitated by using NVivo 12 Plus [22]. Recruitment and retention rates for the study and standard deviations (SDs) of potential primary outcome measures will be estimated and precision summarised using 95%CIs. We will also analyse the within person change over the 24 intervention sessions. All data at an ordinal level will be reported as median values, including data around acceptability (IMI, PACES). All continuous data evaluating preliminary effectiveness on arm function (KinArm data) will be reported as mean with 95% CI. Due to the nature of the feasibility study, it was decided not to conduct any efficacy statistical tests but to use descriptive statistics.A qualitative approach will be used for free text response questions (table 3), with qualitative data analysed using Framework Analysis [20]. Data management will be facilitated by using NVivo 12 Plus [21].

The decision to continue a large follow up trial will dependent on the feasibility measures, which include participant safety and acceptability of this intervention, as well as the logistics of implementing this trial. We have set criteria to be met in order to progress this study further.

## Data Availability

All data produced in the present work are contained in the manuscript

## Data analysis summary

The premise of this study is to test the feasibility of a high-dose neuroanimation intervention in people with MS. The comprehensive feasibility study design will allow to gather vital information on the process, resource, management, acceptability, and scientific feasibility of our intervention. Moreover, the design will allow for a sample size calculation for a future RCT by collecting outcome measure data on motor and cognitive function.

## Ethics and dissemination

Ethics approval for this study has been obtained from the ECU Human Research Ethics Committee (2020-01763-vandergroen). Dissemination of study findings is planned for peerreviewed journals, (inter)national conferences and associated media releases.

## Contributors

OvdG is the principal investigator. OvdG, DE, YL and JS conceived and designed the study. OvdG wrote the first draft of the manuscript with subsequent revisions from all authors. OvdG will lead the study operations with support of the research team.

