## Supplement 1 for "Motor Improvement in Neurological Conditions (MINC): Multiple Sclerosis Design and methods of a single-arm feasibility study"

**Supplement 1 Detailed description of outcome measures**

***Qualitative questions***

A member of the research team not involved in the intervention delivery will ask the participants the questions after the training intervention. If the participants indicates that they are very fatigued during the assessment, they will have the option to conduct the interview via videoconference. Interviews will be recorded and transcribed.

***Heart-rate (HR) data***

HR is recorded during each session and will be used to describe exercise intensity. Predicted HR max will be calculated and the HR value will be expressed as percentage of HRmax.

***Depression, Anxiety and Stress Scale (DASS-21)***

The DASS-21 will be used to assess anxiety, depression and stress symptomatology. Participants will be instructed to indicate their response to each of the 21 items pertaining to depression, anxiety and stress using a four-point Likert scale. Scores will then be used to categorise the participant as normal, mild, moderate, severe or extremely severe for each component.

***Symbol digits modality test***

In this test, a measure of processing speed, a coding key with nine abstract symbols is presented and each paired with a number between 1 and 9. Participants are asked to indicate which numbers go with symbols that are presented in a long string. The participant is given 120seconds to report as many numbers that go with the corresponding symbols.

***KinArm cognitive test Spatial span***

The spatial span test will be used to assess memory and attention. Participants will be asked to repeat a sequence of events displayed on the KinArm robot.

***KinArm cognitive test Trail making***

The trail making test will be used to assess visual attention and task switching. Participants will be asked to complete two different forms: Form A involves drawing a line connecting circles in numerical order, while form B requires participants to switch between numerical and alphabetical order when connecting the circles. This test assesses visual attention as well as task switching.

***Action research arm test (ARAT)***

The ARAT is an evaluative measure to assess specific changes in upper limb function. It assesses a patient’s ability to handle objects differing in size, weight and shape and therefore can be considered to be an arm-specific measure of activity limitation

***Fatigue Severity Scale* *(FSS)***

The fatigue severity scale is a 9-item questionnaire assessing the severity of fatigue and its effect on daily activities and lifestyle. Participants will be instructed to indicate how fatigue has affected their usual activities in the last week using a seven-point Likert scale. A greater score on the fatigue severity scale is indicative of a higher level of fatigue.

***Fatigue Visual Analogue Scale (VAS)***

On this scale participants indicate their current level of fatigue, on a scale of 0 – 10, with 0 being no fatigue and 10 maximal fatigue.

***Rate of Perceived Exertion (RPE)***

The RPE will be determined after each training session with a rating on the Borg scale.

***Timed-up-and go***

This test determines falls risk and measure the progress of balance, sit to stand and walking. The patient sits on a chair and on a ‘Go’ signal she gets up, walks 3m, turns around and sits down again.

***Functional walk test* (10m walk test)**

This is a measure of self-paced walking ability and functional capacity.

***KinArm Measures***

Quantitative assessment of movement will be performed using the KINARM exoskeleton robot (BKIN Technologies Ltd., Kingston, ON, Canada). The KINARM uses advanced robotics technology to quantify human voluntary motor control through flexion and extension movements at the shoulder and elbow joints. Participants will be seated in the KINARM exoskeleton robot which allows 2D horizontal planar arm movement. A virtual reality system will be used to display spatial objects in the horizontal workspace of the arms. A screen will obscure the visual feedback of the hands, in order to make patients rely on fingertip position feedback which will be provided on the display. Preprogramed standardized tasks will be described immediately prior to being performed, and will be administered by experienced operators. This device has been clinically validated to study upper limb motor behaviour.

***Activities Balance Confidence Scale***

This is a self-reported measure of balance confidence in performing various activities without losing balance or experiencing a sense of unsteadiness.

***MSWS-12 (MS walking scale 12 self report)***

This is a self-report measure of the impact of MS on the individuals walking ability.

*Leeds MS Quality of life scale* This scale assesses quality-of-life in people with MS. It consist of 8 groups of statements made by people living with MS and the person indicates how much he/she agrees with the statements.

***Leeds MS Quality of Life***

This scale assesses quality-of-life in people with MS. It consist of 8 groups of statements made by people living with MS and the person indicates how much he/she agrees with the statements.

***Saliva/Genetic factors***

As part of the SPIN research program, before the start of the intervention, on the first training day, a saliva sample will be collected for DNA and RNA analysis. At the start of follow up visit 1 another RNA saliva sample will be collected. These data will be stored for future analysis within the SPIN research program. The SPIN research program will be conducting an exploratory analysis of this DNA. This means that we will be looking for new (novel) genetic information, the clinical significance of which is not yet known.

***Intrinsic Motivation Inventory (IMI)***

The intrinsic motivation inventory is a multidimensional questionnaire structured in different subscales. In the current study we use five subscales: “Interest/Enjoyment,” “Perceived Competence,” “Effort/Importance,” “Pressure/Tension,” “Value/Usefulness”. The interest/enjoyment subscale is considered the self-report measure of intrinsic motivation. Items are rated on seven-point Likert scales (1 “not at all true” to 7 “very true”). Higher scores relate to higher internal motivation, except for the pressure/tension subscale, where higher scores indicate greater feelings of pressure, which is conceived a negative predictor of intrinsic motivation.

***Physical Activity Enjoyment Scale (PACES)***

The PACES is an 18-item self-administered scale assessing the enjoyment of a physical activity. The PACES uses a seven-point semantic differential approach. The total score is calculated so that a higher score indicates a higher level of enjoyment.

***Godin Exercise Leisure Time Questionnaire***

This is a common self-report measure of physical activity for persons with multiple sclerosis.

***Visual analogue motivation towards exercise***

This is a scale from 1 to 10 in which participants indicate how motivated they are to engage in the exercise.

***Exercise Self-efficacy***

This list of statements is designed to assess someone’s beliefs in their ability to continue exercising on a three time per week basis at moderate intensities (upper end of your perceived exertion range), for 40+ minutes per session in the future.
