## Supplement 2 for "Motor Improvement in Neurological Conditions (MINC): Multiple Sclerosis Design and methods of a single-arm feasibility study"

**Mapping of evaluation plan for aim 1**

| **Respondent** | **Questioning** | **Response style** | **Domain** |
| --- | --- | --- | --- |
| Intervention participants | Please describe in detail the barriers for you to participate in this study | **Free text** | **Process** |
|  | What did you think of the length of the testing? | **0 – 5 Likert scale plus free text** | **Scientific** |
|  | Please describe in detail the positive outcomes you have experienced from playing the dolphin game (MINC MS)? | **Free text** | **Scientific** |
|  | Please describe in detail why you think you have experienced these positive outcomes from the dolphin game (MINC MS)? | **Free text** | **Scientific** |
|  | Please describe in detail what circumstances might have made you unable to experience these positive outcomes. | **Free text** | **Scientific** |
|  | What did you think of the dolphin game (MINC: MS) intervention duration? | **Free text** | **Scientific** |
|  | How suitable was the dolphin game (MINC: MS) program to your MS symptoms? | **0–5 Likert scale** | **Scientific** |
|  | How suitable was the dolphin game (MINC:MS) program to your personal fitness level? | **0–5 Likert scale** | **Scientific** |
|  | How much would you recommend the dolphin game (MINC: MS) program to others like you? | **0–5 Likert scale** | **Scientific** |
|  | Please describe in detail in what circumstances (to who, for when, for what) would you recommend the dolphin game (MINC:MS) to others like you. | **Free text** | **Scientific** |
|  | Please describe in detail in what circumstances (to who, for when, for what) would you not recommend the dolphin game (MINC:MS) to others like you | **Free text** | **Scientific** |
|  | Please describe in detail how satisfied were you with the overall dolphin game (MINC:MS) program? | **Free text** | **Acceptability** |
|  | Please describe in detail how satisfied you were with: The exercises you had to do? Why do you think that? | **Free text and 0–5 Likert scale** | **Acceptability** |
|  | How many days per week would you like to do this intervention? | **Free text** | **Acceptability** |
|  | How did being part of the dolphin game (MINC: MS) make you feel? (Please explain your response and provide examples) | **Free text** | **Acceptability** |
|  | How convenient was using the dolphin game (MINC: MS) as a form of rehabilitation?  (Please explain your response and provide examples) | **Free text and 0–5 Likert scale** | **Acceptability** |
|  | Did you think being offered the dolphin game (MINC: MS) was appropriate for your condition?  (Please explain your response and provide examples) | **Free text and 0–5 Likert scale** | **Acceptability** |
|  | How much of a priority is your rehabilitation in your day-to-day life? (Please explain your response and provide examples) | **Free text and 0–5 Likert scale** | **Acceptability** |
|  | Did you understand the overall aim of the MINC study?  (Please explain your response and provide examples) | **Free text and 0–5 Likert scale** | **Acceptability** |
|  | How good was the dolphin game (MINC: MS) at meeting your needs? (Please explain your response and provide examples) | **Free text and 0–5 Likert scale** | **Acceptability** |
|  | Did you feel able to do what you needed to in order to perform the tasks in the dolphin game (MINC: MS)? (Please explain your response and provide examples) | **Free text and 0–5 Likert scale** | **Acceptability** |
| **Staff delivering the intervention/ Clients primary therapist** | Please describe in detail what positive outcomes did your clients experience from engaging with the dolphin game (MINC:MS)? | **Free text** | **Scientific** |
|  | Please describe in detail why do you think they have these positive outcomes from the dolphin game (MINC:MS)? | **Free text** | **Scientific** |
|  | Please describe in detail what circumstances might have made them unable to experience these positive outcomes. | **Free text** | **Scientific** |
|  | How suitable was the dolphin game (MINC: MS) program to the symptoms associated with MS? | **0–5 Likert scale** | **Scientific** |
|  | How much would you recommend the dolphin game (MINC:MS) program to other clinicians? | **0–5 Likert scale** | **Scientific** |
|  | Please describe in detail what circumstances (to who, for when, for what) would you recommend the dolphin game (MINC:MS) to other clinicians for delivery to persons with MS? | **Free text** | **Scientific** |
|  | Please describe in detail what circumstances (to who, for when, for what) would you not recommend the dolphin game (MINC:MS) to other clinician for delivery to persons with MS? | **Free text** | **Scientific** |
|  | Please describe in detail how satisfied do you think your clients were with: The overall dolphin game (MINC:MS) program?  Please explain your response. | **Free text and 0–5 Likert scale** | **Acceptability** |
|  | Please describe in detail how satisfied do you think your clients were with:  The exercises participants had to do? Why do you think that? | **Free text and 0–5 Likert scale** | **Acceptability** |
|  | How do you feel about delivering the dolphin game (MINC: MS)?  (Please explain your response and provide examples) | **Free text** | **Acceptability** |
|  | How convenient was delivering the dolphin game (MINC: MS) as a form of rehabilitation?  (Please explain your response and provide examples) | **Free text and 0–5 Likert scale** | **Management** |
|  | Did you think delivering the dolphin game (MINC: MS) was appropriate for individuals with MS? (Please explain your response and provide examples) | **Free text and 0–5 Likert scale** | **Acceptability** |
|  | How much of a priority is your client’s rehabilitation in your day-to-day life? (Please explain your response and provide examples | **Free text and 0–5 Likert scale** | **Acceptability** |
|  | What did you understand about the MINC: MS intervention and what it involved in delivering it? (Please explain your response and provide examples) | **Free text and 0–5 Likert scale** | **Acceptability** |
|  | How good do you think the dolphin game (MINC: MS) is at meeting client’s needs? (Please explain your response and provide examples) | **Free text and 0–5 Likert scale** | **Acceptability** |
|  | Did you feel able to do what you needed to in order to deliver the tasks in the dolphin game (MINC: MS)? (Please explain your response and provide examples) | **Free text and 0–5 Likert scale** | **Acceptability** |
| **All staff** | How much time in minutes, did you spend on recruitment ? | **Free text** | **Management** |
|  | Please note down the time you spend on data collection in minutes, including time for outcome assessments. | **Free text** | **Management** |
|  | Please describe in detail what challenges did you experience for yourself during the intervention or assessments? (e.g. training needs?) | **Free text** | **Management** |
